# Non-severe SARS-CoV-2 infection is characterised by very early T cell proliferation independent of type 1 interferon responses and distinct from other acute respiratory viruses

**DOI:** 10.1101/2021.03.30.21254540

**Authors:** Aneesh Chandran, Joshua Rosenheim, Gayathrie Nageswaran, Leo Swadling, Gabriele Pollara, Rishi K Gupta, Jose Afonso Guerra-Assuncao, Annemarie Woolston, Tahel Ronel, Corrina Pade, Joseph Gibbons, Blanca Sanz-Magallon Duque De Estrada, Marc Robert de Massy, Matthew Whelan, Amanda Semper, Tim Brooks, Daniel M Altmann, Rosemary J Boyton, Áine McKnight, Charlotte Manisty, Thomas Alexander Treibel, James Moon, Gillian S Tomlinson, Mala K Maini, Benjamin M Chain, Mahdad Noursadeghi, COVIDsortium Investigators

## Abstract

The correlates of natural protective immunity to SARS-CoV-2 in the majority who experience asymptomatic infection or non-severe disease are not fully characterised, and remain important as new variants emerge. We addressed this question using blood transcriptomics, multiparameter flow cytometry and T cell receptor (TCR) sequencing spanning the time of incident infection. We identified a type 1 interferon (IFN) response common to other acute respiratory viruses, and a cell proliferation response that discriminated SARS-CoV-2 from other viruses. These responses peaked by the time the virus was first detected, and in some preceded virus detection. Cell proliferation was most evident in CD8 T cells and associated with rapid expansion of SARS-CoV-2 reactive TCRs. We found an equally rapid increase in immunoglobulin transcripts, but circulating virus-specific antibodies lagged by 1-2 weeks. Our data support a protective role for rapid induction of type 1 IFN and CD8 T cell responses to SARS-CoV-2.

## Introduction

The host response in non-severe SARS-CoV-2 infection during the first epidemic wave, prior to vaccination, incorporates the mechanisms of effective host-defence in naïve populations. To date, our knowledge has been limited to immune responses after the detection of the virus or onset of symptoms, and to cross-sectional studies in which the time of infection was undefined. As a result, the temporal kinetics and relationships between the earliest immune responses to infection are not known. These early events, among the majority that experience asymptomatic infection or mild disease not requiring hospitalisation, may provide new insights into the determinants of immune protection in naïve populations that may also be relevant to emerging variants that escape vaccine mediated protection. We sought to address this question at systems level by genome-wide transcriptional profiling of weekly blood samples before, during and after incident SARS-CoV-2 infections during the first epidemic wave in London, and compared our findings with responses to other acute respiratory viruses using publicly available data from human challenge experiments.

## Results

### Type 1 interferon and cell proliferation responses from one week before to three weeks after detection of asymptomatic and mild SARS-CoV-2 infection

We undertook a nested case-control study derived from a cohort of 400 healthcare workers at one London hospital recruited from 23^rd^ March 2020 to undergo weekly nasopharyngeal swab PCR tests and blood sampling when fit to attend work, as previously described^1–5^. In this cohort, we detected 45 incident infections by PCR. Among these cases, we obtained 114 blood transcriptional profiles from 41 individuals spanning three weeks before to three weeks after the first PCR positive result, including 12 individuals for whom samples were available before the first positive PCR. We also profiled convalescent samples from 16/41 individuals 5-6 months later. We compared these data to blood transcriptional profiles obtained from baseline samples in 55 sequential uninfected controls who remained PCR and seronegative for SARS-CoV-2 during follow up (Supplementary Figure 1, Supplementary Table 1). None of the individuals who became infected required hospitalisation. Among 38 individuals for whom blood transcriptomic data were available at the time of first positive PCR, 29 had no contemporary symptoms attributable to SARS-CoV-2 infection. Genome-wide transcriptional profiles from those who experienced an infection showed greatest perturbation compared to uninfected controls at the time of the first positive PCR test, independent of symptoms (Figure 1a, Supplementary Figure 3). Their profiles were significantly different from uninfected controls from the week before the first positive PCR to three weeks afterwards. Six month convalescent samples from a subset of these individuals were not significantly different to uninfected controls, indicating that the blood transcriptome had fully reverted to the baseline.

**Figure 1.**
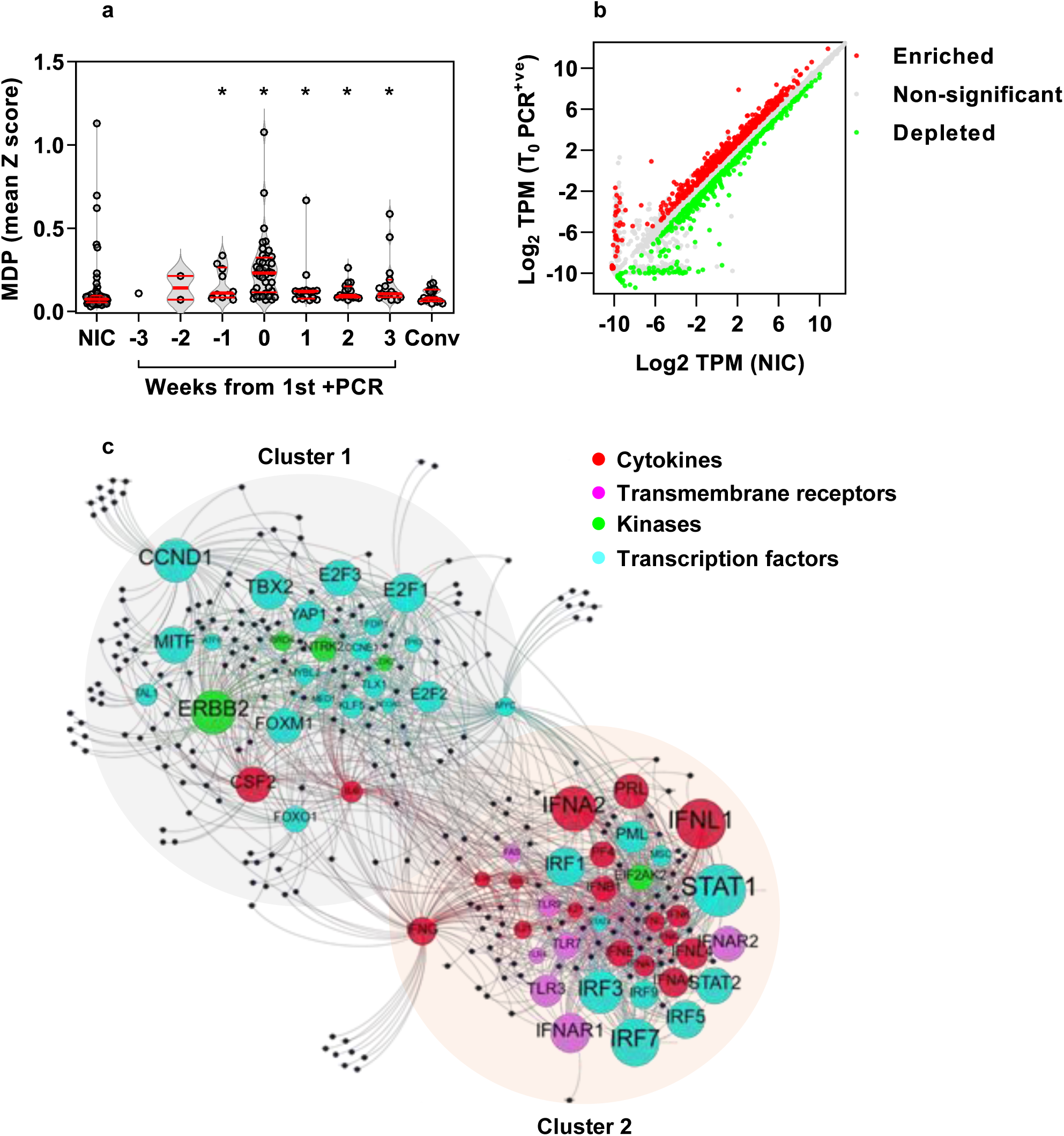
Incident SARS-CoV-2 infection associated with perturbation of blood transcriptome reflecting type 1 interferon and cell proliferation responses. **(A)** Molecular degree of perturbation (MDP) in blood transcriptomes for each individual expressed as the mean of genome-wide standard deviations (Z scores) from the mean of non-infection controls (NIC), among NIC, individuals with incident infection stratified by weeks from first positive PCR and convalescent samples 5-6 months after incident infection. Individual data points shown with violin plots depicting median, IQR and frequency distributions. (*FDR<0.05 by Kruskal-Wallis Test for each group compared to NIC) **(B)** Differentially expressed genes in blood transcriptomes at time of first positive PCR compared to NICs. **(C)** Predicted upstream regulators (labelled nodes) stratified by molecular function for differentially expressed genes (black nodes). Size of the nodes for upstream regulators proportional -Log10 p-value. Nodes clustered using Force Atlas 2 algorithm.

To investigate the host response to infection, we identified differentially expressed transcripts by comparison of profiles from the time of first positive viral PCR, to those of uninfected controls (Figure 1b). These were subjected to upstream regulator enrichment analysis to identify molecular pathways predicted to be activated at the level of cytokines, transmembrane receptors, kinases and transcription factors that may be responsible for differential gene expression (Supplementary File 1). We filtered groups of target genes associated with each upstream regulator to include only those that had significantly greater co-correlated expression than would be expected at random in our blood transcriptomes, in order to increase our confidence that these represent co-regulated genes in a given molecular pathway (Supplementary Figure 4). Among those that were retained, the associated upstream regulators formed two clusters resulting from overlapping associations with target genes (Figure 1c), reflecting two predominant biological pathways. These were type 1 interferon (IFN) responses and cell cycle activity, as surrogates for innate immune activation and cellular proliferation respectively (Supplementary Figure 5a). We collated the differentially expressed genes linked to the most statistically enriched upstream regulator in each of the two clusters as a transcriptional module, resulting in a Signal Transducer and Activator of Transcription (STAT)1-regulated module to represent type 1 IFN responses and a Cyclin D1 (CCND1)-regulated module to represent the cell proliferation response. The validity of the functional annotation for each of these modules was confirmed by investigating their correlation and covariance with independently derived transcriptional signatures for type 1 and type 2 IFN responses, and for cell proliferation. The STAT1 module correlated with both IFN response modules, but showed much greater covariance with the type 1 IFN signatures (Supplementary Figure 5b), consistent with our bioinformatic analysis of the functional pathway represented by this cluster of differentially expressed genes. Similarly, we found that the CCND1 regulated gene expression module showed good correlation and covariance with an independently derived cell proliferation module (Supplementary Figure 5c). Type 1 IFN and cell proliferation responses both peaked with co-incident infection (Figure 2a-b), but significant increases in these responses were also evident in the week before the first positive PCR result. Type 1 IFN responses remained significantly elevated for one week after the first positive PCR, whereas the cell proliferation response remained elevated for two weeks after the first positive PCR. The peak of each of these responses over this time course, discriminated infected individuals from non-infected controls with area under the receiver operating characteristic curve (AUROC) of 0.87 (95% confidence interval 0.78-0.94) and 0.92 (0.87-0.98) for the STAT1 and CCND1-regulated modules respectively (Supplementary Figure 6a), giving a measure of the consistency of both these responses in infected individuals. Despite this and the overlap in the temporal profiles of these two responses, the enrichment of STAT1 and CCND1-regulated modules representing each response at the individual participant level, did not correlate, suggesting that they may be independently regulated or subject to idiosyncratic capacity for each of these responses at the level of individual participants (Figure 2c). The same observation was evident for differentially expressed genes combined as modules associated with each of the upstream regulators that reflected type 1 IFN or cell proliferation modules (Figure 2d).

**Figure 2.**
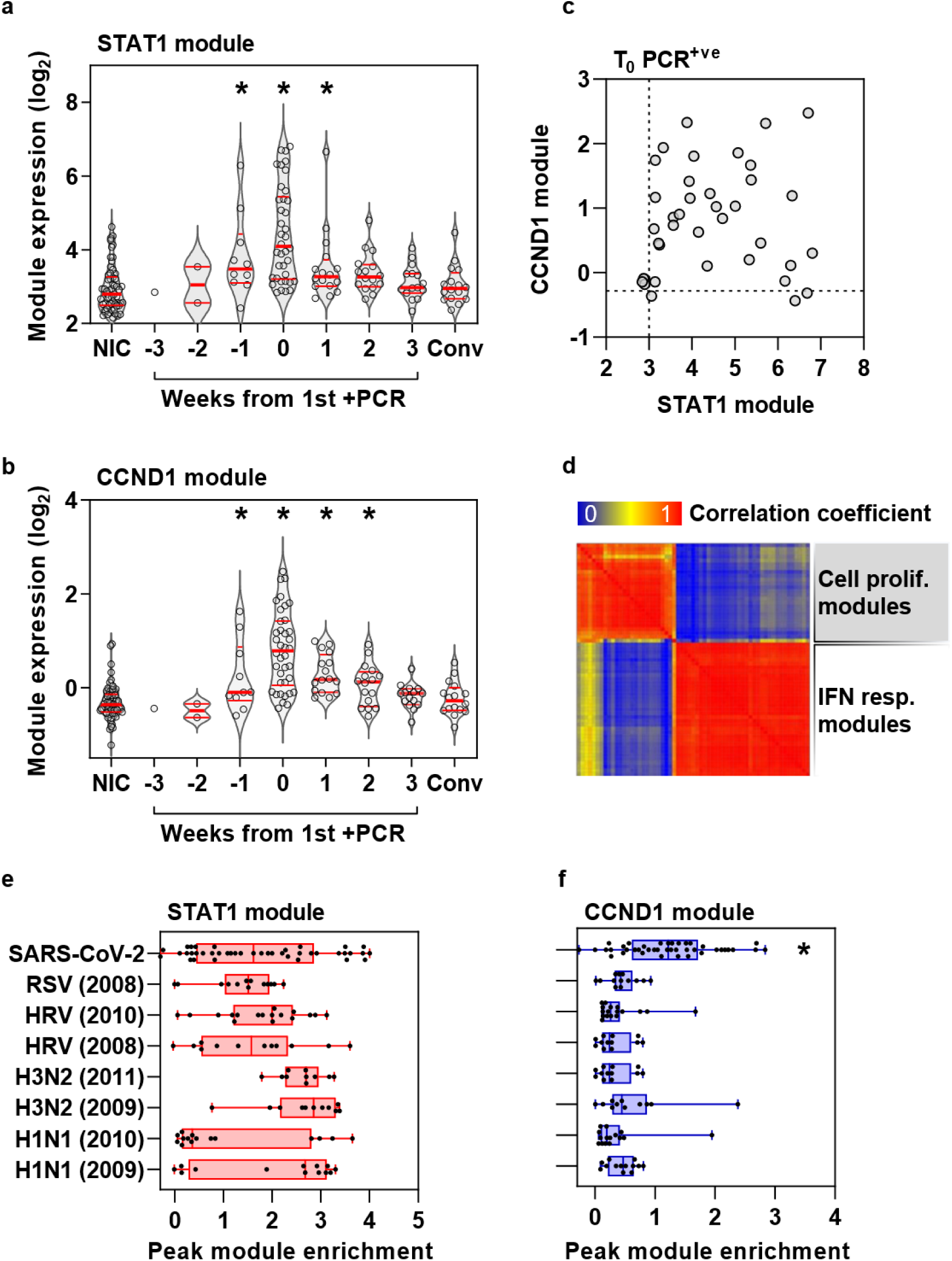
Cell proliferation response discriminates SARS-CoV2 infection from other acute viruses and is not correlated to type 1 IFN response. **(A)** Expression of STAT1 module (representative of type 1 IFN response) and **(B)** CCND1 module (representative of cell proliferation response) in blood transcriptomic data stratified by time to first positive SARS-CoV-2 infection, compared to non-infection controls (NIC) and convalescent samples 5-6 months after incident infection. Individual data points shown with violin plots depicting median, IQR and frequency distributions. (*FDR<0.05 by Kruskal-Wallis Test for each group compared to NIC). **(C)** Comparison of STAT1 and CCND1 module expression at time of first positive PCR (dashed lines represent the upper limit of the 95% confidence interval of median of NIC), and **(D)** co-correlation matrix between all type 1 IFN and cell proliferation modules at time of first positive PCR. **(E)** Comparison of STAT1 and **(F)** CCND1 module expression associated with co-incident SARS-CoV-2 infection compared to peak expression of these modules in experimental human challenge infections using Respiratory Syncytial Virus (RSV), Human Rhinovirus (HRV) or Influenza virus (H3N2 and H1N1), stratified by different data sets indicated by year. (*FDR<0.05 by Kruskal-Wallis Test in SARS-CoV-2 infection compared to all other groups).

### Cell proliferation responses distinguish SARS-CoV-2 infection from other acute respiratory viruses and are predominantly attributable to T cell responses

Next, we compared the type 1 IFN and cell proliferation response to incident SARS-CoV-2 infection with those of other acute respiratory viruses, by comparing the peak expression of the STAT1 and CCND1-regulated modules in our cohort to that of publicly available longitudinal blood transcriptomic data derived from human challenge experiments with Respiratory Syncytial Virus (RSV), Human Rhinovirus (HRV) and Influenza viruses (Supplementary Figure 6b)^6^. Comparable enrichment of the type 1 IFN response was evident in each of these infections (Figure 2e), but the cell proliferation response was significantly greater to SARS-CoV-2 than the peak response to any of the other acute respiratory virus infections (Figure 2f). We tested the hypothesis that the cellular proliferation response may arise from rapid B cell or T cell expansion in response to infection by evaluating the correlation between the CCND1 module and expression of validated cell-type specific signatures (Figure 3a). CCND1 module expression correlated with the transcriptional signature for T cells, but not B cells. The relationship between the cell proliferation response and T cell subsets was stronger for the CD8 T cell signature than for the CD4 T cell signature. To corroborate these findings, we undertook multiparameter flow cytometry of peripheral blood mononuclear cells (PBMC) obtained in a subset of participants with contemporaneous PCR positive infection and compared these to PBMC from uninfected controls (Supplementary Figure 7). Representation of the pooled multiparameter flow cytometry data by tSNE incorporating all T cells revealed new populations of CD4 and CD8 positive cells in samples from infected participants, which also exhibited the highest levels of Ki67 staining as a marker of cell proliferation (Figure 3b), and accompanied by HLA-DR expression as a marker of cell activation in exemplar cases (Figure 3c). In this subset of samples only Ki67 staining of CD8 T cells was statistically enriched in infected individuals compared to controls (Figure 3d). Of all other lymphocyte subsets only NKT cells showed a significant increase in Ki67 positive staining (Supplementary Figure 8a), but comprised on average 3% of circulating lymphocytes compared to T cells that comprised approximately 40% (Supplementary Figure 8b).

**Figure 3.**
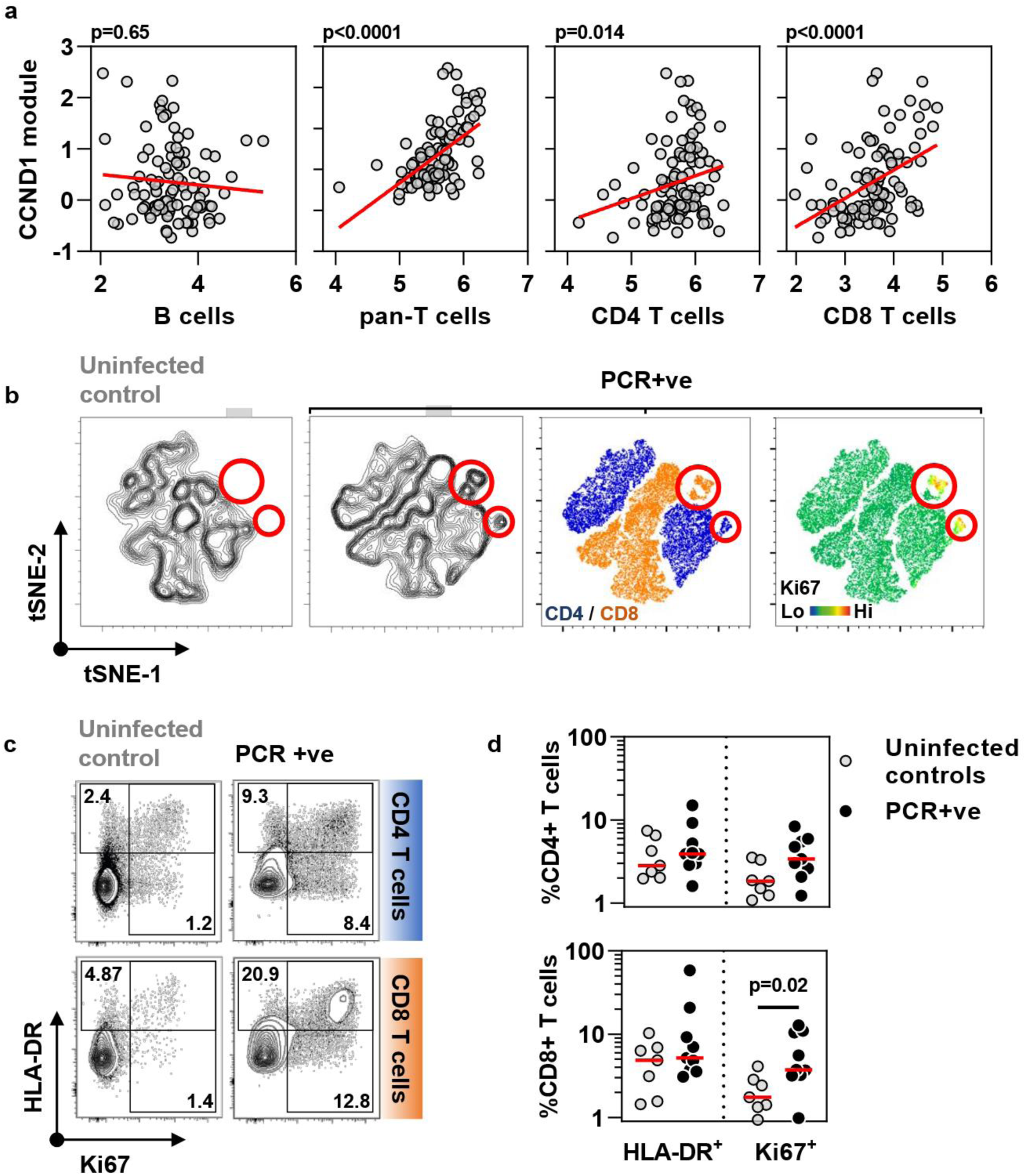
Cell proliferation response to SARS-CoV-2 infection in blood transcriptomic data is attributable to T cell proliferation. **(A)** Correlation of CCND1 module (representative of cell proliferation response) in all time points (−3 to +3 weeks) from individuals with SARS-CoV-2 infection with each of blood transcriptomic modules representative of B cells, pan-T cells, CD4 T cells and CD8 T cells (regression lines shown in red, p values for Spearman rank correlations). **(B)** Representative tSNE plots from one non-infected control and one individual with co-incident PCR positive SARS-CoV-2 infection derived from multiparametric flowcytometry data of CD4 and CD8 positive T cells. Contour plots are shown in the two left hand panels, followed by dot plots stratified by CD4/CD8 staining and relative Ki67 staining as a proliferation marker. Red circles highlight a population of Ki67 high CD4 and CD8 cells exclusive to the PCR+ group. **(C)** Representative flow cytometry data for HLA-DR and Ki67 staining in either CD4 T cells or CD8 T cells from one non-infected control and one individual with co-incident PCR positive SARS-CoV-2 infection. Numbers indicate % positive for each marker including double positives **(D)** Summary HLA-DR and Ki67 staining data from seven uninfected controls and nine individuals with coincident infection in either CD4 T cells or CD8 T cells. P value shown for Mann Whitney Test.

### Rapid clonal T cell expansion in response to SARS-CoV-2 infection associated with significant enrichment of SARS-CoV-2 reactive T cell receptors

To further evaluate the rapid T cell response to SARS-CoV-2 infection, we undertook sequencing of TCR alpha and beta chains in longitudinal samples to reflect dynamic changes in the T cell clonal repertoire. An expanded clone will increase or decrease in frequency depending on the sampling time point before and after the peak response. Therefore, we identified expanded TCR sequences as being statistically enriched at one time point compared to at least one other time point, and summed the total number of expanded sequences for these TCRs at each time point per million of total TCR sequences (Supplementary Figure 9). These were compared to expanded sequences identified in the same way among a subset of six uninfected controls in whom we undertook TCR sequencing in samples from five successive weeks. By comparison to the pooled data from controls, a significant increase in expanded TCRs was evident in infected individuals by the time of the first positive PCR test up to a maximum abundance of >6% of total T cell sequences, and persisted for at least three weeks for both alpha and beta chains (Figure 4a, Supplementary Figure 10a). The abundance of expanded TCR sequences correlated significantly with the CCND1 but not STAT1 regulated module, consistent with the hypothesis that the proliferation response reflected expansion of T cell clones (Figure 4b-c).

**Figure 4.**
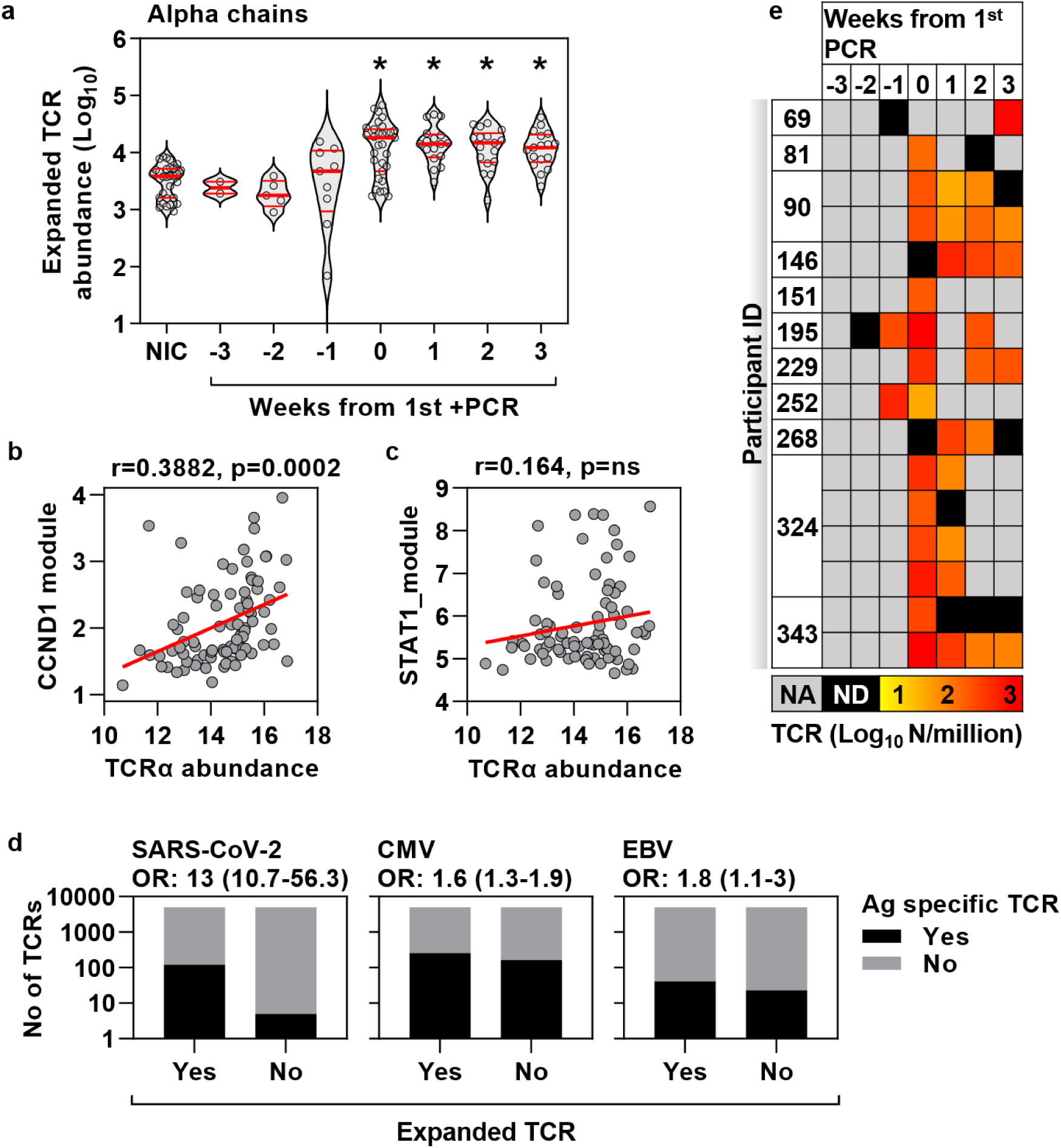
Cell proliferation response to co-incident SARS-CoV-2 infection associated with expansion of TCR clones enriched for SARS-CoV-2 reactive TCRs. **(A)** Enumeration of expanded TCR alpha chain abundance (per million total sequences) in non-infection controls and samples from infected individuals stratified by time from first positive PCR. Individual data points shown with violin plots depicting median, IQR and frequency distributions. (*FDR<0.05 by Kruskal-Wallis Test for each group compared to NIC). **(B)** Correlation of CCND1 module and **(C)** STAT1 module with TCR alpha-chain sequences (Log_2_ per million sequences). Regression lines shown in red, with r and p values for Spearman rank correlations. **(D)** Number of antigen-specific TCR sequences (alpha and beta-chains) for SARS-CoV-2, cytomegalovirus (CMV) and Epstein-Barr Virus (EBV) among expanded TCR sequences in all time points (−3 to +3 weeks) from individuals with SARS-CoV-2 infection and among non-expanded TCRs from the same samples, giving the odds ratio (±95% confidence interval, Fisher’s exact test) for enrichment of antigen specific TCR sequences in each case. **(E)** Frequency heat map of individual (rows) SARS-CoV-2 reactive TCRs (alpha and beta-chains) identified among expanded TCR sequences in all time points (−3 to +3 weeks) from individuals with SARS-CoV-2 infection. NA=no sample available; ND=not detected in sample).

T cell clonal expansion was not explained by changes in MAIT cell or NKT cell-associated TCR sequences (Supplementary Figure 10b). Comparison with sequence data for known antigen reactive TCRs in VDJdb^7^ confirmed that expanded TCRs among infected individuals were most highly enriched for SARS-CoV-2 reactive T cells, compared to CMV or EBV reactive TCRs to represent non-specific bystander T cell proliferation (Figure 4d). Identifiable SARS-CoV-2 reactive TCR sequences available in VDJdb were evident in 11 individuals (Supplementary File 2). In all but two of these individuals, expanded SARS-CoV-2 reactive TCRs were present by the time of 1^st^ positive PCR test (Figure 4e).

### Circulating virus specific antibodies lad two weeks behind transient increase in immunoglobulin gene expression in response to SARS-CoV-2 infection

The finding that the CCND1-regulated module did not correlate with our B cell signature does not exclude a B cell response. Emergence of antibodies to SARS-CoV-2 has been reported as early as early as five days after symptom onset^8^. We found increased expression of immunoglobulin (Ig) constant heavy and light chain transcripts, which peaked at the time of first PCR virus detection but was evident from one week before to two weeks after first PCR detection (Figure 5a-b). The increase in Ig gene expression in blood was less sustained than TCR expansion, and returned to baseline by three weeks after the first positive PCR. In contrast, circulating antibodies to SARS-CoV-2 S1 spike protein that correlate with virus neutralisation were not detectable until one week after the incident infection (Figure 5c) and continued to increase in this cohort for eight weeks^3,4^.

**Figure 5.**
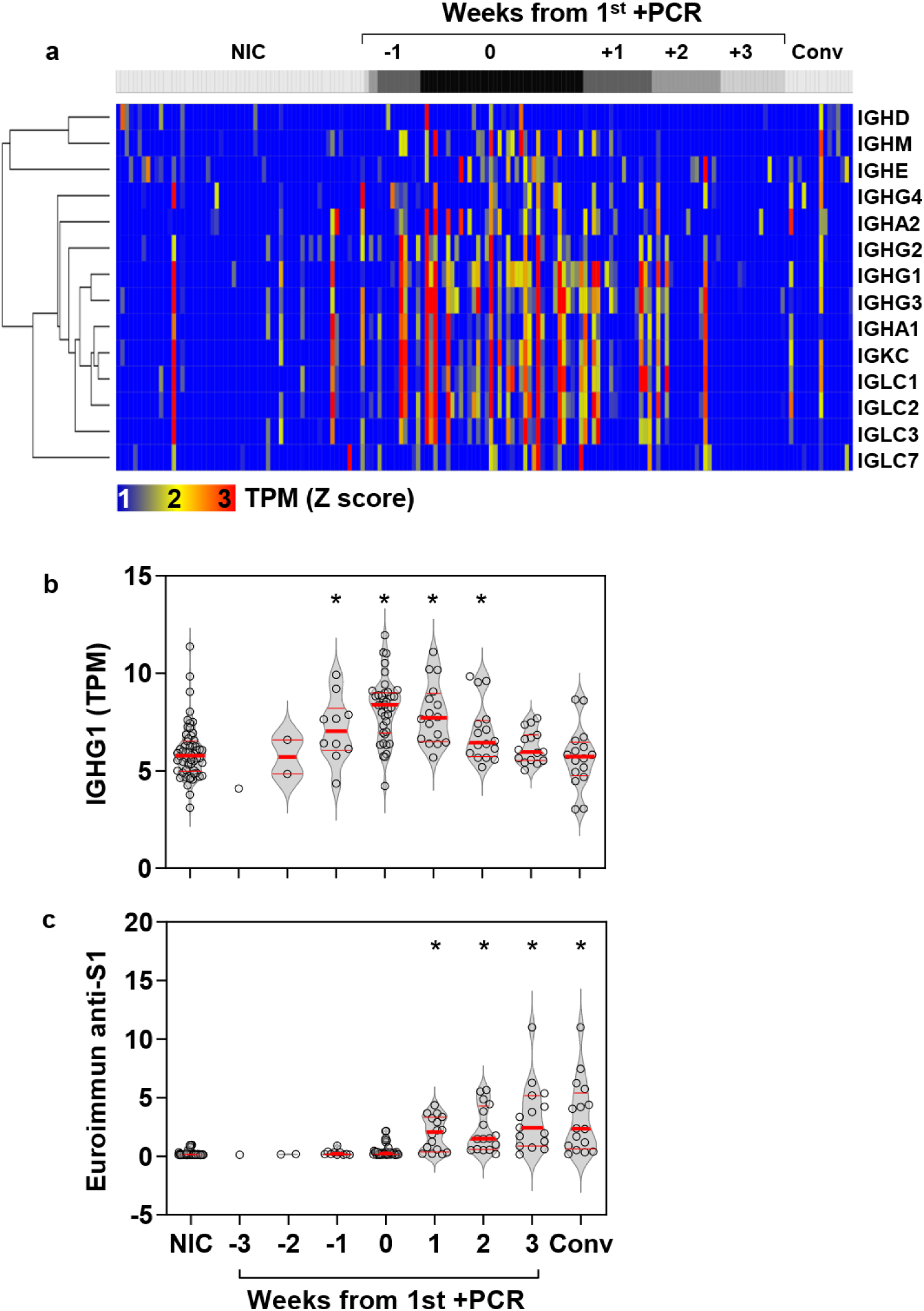
Enriched immunoglobulin gene expression and antibody response to incident SARS-CoV-2 infection. **(A)** Heatmap of immunoglobulin constant heavy and light chain gene expression in blood per individual (columns) stratified by time to first positive SARS-CoV-2 infection, compared to non-infection controls (NIC) and convalescent (Conv) samples 5-6 months after incident infection, presented as standardized (Z) scores of transcripts per million (TPM) using mean and standard deviation of NIC. **(B)** Blood TPM of IGHG1 and **(C)** relative IgG anti-S1 antibody levels stratified by time to first positive SARS-CoV-2 infection, compared to NIC and Conv samples. Individual data points shown with violin plots depicting median, IQR and frequency distributions. (*FDR<0.05 by Kruskal-Wallis Test for each group compared to NIC).

## Discussion

To the best of our knowledge, we report the earliest *in vivo* immune responses to SARS-CoV-2 infection available to date, enabled by serial sampling of individuals at risk of infection during the peak of the first epidemic wave in London. The general paradigm for early antiviral host defence is dominated by induction of type 1 IFNs. Attenuated responses as a result of autoantibodies to type 1 IFNs, and genetic polymorphisms associated with reduced expression of a type 1 IFN receptor subunit or with reduced expression of the IFN-inducible oligoadenylate synthetase (OAS) gene cluster have all been associated with severe disease^9,10^. These provide strong evidence that type 1 IFN responses contribute to effective protection against SARS-CoV-2 infection. We show that type 1 IFN responses can precede PCR detection of the virus and therefore may exert their protective effects in the earliest phases of infection, independent of symptoms. We propose that such early detection of IFN-inducible genes in the blood transcriptome may arise from localised immune responses as a result of leukocyte trafficking through lymphoid tissues or the site of infection, and may provide greater sensitivity than detection of circulating IFNs. As we have previously reported, an additional translational application of this finding is the detection of IFN-inducible transcripts in blood, as a diagnostic biomarker of early viral infection that may precede PCR detection of the virus and symptoms^11^.

Alongside type 1 IFN responses, we detected an early cell proliferation response in the blood transcriptome, which we primarily attribute to CD8, and to a lesser extent CD4 T cell proliferation by correlation with cell-type specific transcriptional modules, corroborated by flow cytometry to show significant increase in Ki67 positive CD8 T cells and TCR sequencing to show expansion of T cell clones. Whilst type 1 IFN responses were evident in a range of other acute respiratory virus infections modelled in human challenge experiments^6^, the early T cell response to SARS-CoV-2 in our study was significantly greater than in other viral infections. By comparison with emerging databases of SARS-CoV-2 specific TCRs in VDJdb, we were able to show that expanded T cell clones were most enriched for SARS-CoV-2 reactive cells, and that these were already evident by the time of first positive virus PCR. In individuals with COVID-19, T cell reactivity has been reported as early as 5-10 days after the onset of symptoms^12^. Importantly, in one report, T cell proliferative responses to SARS-CoV-2 were evident in 92% of family contacts of COVID-19 cases independently of serostatus^13^, and some people may have pre-existing cross-reactive T cells arising from previous seasonal coronavirus exposure^13–18^. These may be expected to contribute to early viral clearance, analogous to findings in infuenza^19–21^. If this were the primary driver of rapid T cell responses to SARS-CoV-2 infection, the fact that the early proliferative response discriminated infected and uninfected individuals with an AUROC of 0.92 would require pre-existing T cell priming to be a near ubiquitous feature of asymptomatic or non-severe infection. Consistent with this hypothesis, among the largest studies of pre-pandemic blood samples, heterologous T cell reactivity to SARS-CoV-2 peptides with proven similarity to those of pre-existing seasonal coronaviruses has been reported in 81%^18^. In this context, we hypothesise that the variation in T cell proliferative response and the lack of its correlation with type 1 IFN responses may be explained by differential levels of T cell priming in individual participants. We also identified a similarly rapid B cell response represented by transient enrichment of Ig gene expression in blood. We interpret this to represent the transit of activated antigen specific B cells from lymphoid tissues to the predominant site of antibody production in the bone marrow and spleen. Since protective anti-S1 antibodies only became detectable after a two-week lag, we hypothesise that the B cell response may have had a less important role in rapid viral clearance in asymptomatic and non-severe infection.

Our study has some important limitations. The precise time of exposure to SARS-CoV-2 or transmission of infection was not possible to determine. This was offset by including longitudinal samples in 12 subjects before detection of incident infection by PCR, providing enough statistical power to show that both type 1 IFN and cell proliferation responses were statistically enriched in the week before the first positive PCR result. We had limited access to PBMC to assess frequency, phenotypic and functional characteristics of SARS-CoV-2 reactive T cells. The accumulating database of SARS-CoV-2 specific TCR sequences allowed us to relate clonal T cell expansion with antigen-specificity. This only accounted for an extremely small fraction of expanded sequences and does not exclude proliferation of bystander T cells. However, substantially lower levels of enrichment of CMV or EBV specific TCRs among expanded clones, and the lack of enrichment for IFNγ activity or other signatures of T cell activation in the blood transcriptome argue against generalised bystander T cell activation. Future re-analysis of the data (as more sequences across a wider range of HLA haplotypes are reported) will be necessary to evaluate whether the majority of expanded TCRs are ultimately found to recognise SARS-CoV-2. The focus on the blood compartment meant that we do not have direct measurements of responses at the site of host-pathogen interactions. Analysis of bulk RNA samples for transcriptional profiling and TCR sequencing restricted our ability to evaluate transcriptional heterogeneity at the cellular level, further characterise expanded T cell clones or undertake TCR analysis with paired alpha/beta chains. Most importantly, since less than 5% of infections lead to hospitalisation^22^, our study design precluded comparison of severe and non-severe outcomes that would require substantially greater sample size. Nonetheless, our data reflect immune responses in asymptomatic and non-severe infection, which incorporate correlates of effective host defence to natural infection in a naïve population, providing further evidence for the importance of early type 1 IFN and T cell responses. Human challenge experiments that control for variation in time and dose of exposure will offer the best opportunities to acquire the granular detail of early immune responses. Larger scale studies will be required to asses frequency of SARS-CoV-2 T cell reactivity in naïve populations, and determine whether early type 1 IFN or T cell responses predict outcomes. Although vaccine-roll out is likely to be the primary immunological strategy to control the pandemic^23^, understanding the determinants of effective natural immunity will remain a critical objective to enable risk stratification and novel vaccine design as the virus evolves. In particular, identification of the antigenic determinants of the earliest T cell responses in asymptomatic SARS-CoV-2 infection is a priority to inform development of potential universal coronavirus vaccines.

## Methods

### Ethical approval

The study was approved by a UK Research Ethics Committee (South Central - Oxford A Research Ethics Committee, reference 20/SC/0149). All participants provided written informed consent.

### Study design

We undertook a case control study nested within our COVIDsortium health care worker cohort. Participant screening, study design, sample collection, and sample processing have been described in detail previously^24–26^ and the study is registered at ClinicalTrials.gov (NCT04318314). Briefly, healthcare workers were recruited at St Bartholomew’s Hospital, London, UK in the week of lockdown in the United Kingdom (between 23^rd^ and 31^st^ March 2020). Participants underwent weekly evaluation using a questionnaire and biological sample collection (including serological assays) for up to 16 weeks when fit to attend work at each visit, with further follow up samples collected at 6 months. Participants with available blood RNA samples who had PCR-confirmed SARS-CoV-2 infection (Roche cobas® diagnostic test platform) at any time point were included as ‘cases’. A subset of consecutively recruited participants without evidence of SARS-CoV-2 infection on nasopharyngeal swabs and who remained seronegative by both Euroimmun antiS1 spike protein and Roche anti-nucleocapsid protein throughout follow-up were included as uninfected controls.

### Blood RNA sequencing

For ‘cases’, we included all available RNA samples, including convalescent samples at week 24 of follow-up for a subset of participants. For uninfected controls, we included baseline samples only. Genome wide mRNA sequencing was performed as previously described,^27^ resulting in a median of 26 million (range, 19·8–32·4 million) 41 bp paired-end reads per sample. RNAseq data were mapped to the reference transcriptome (Ensembl Human GRCh38 release 100) using Kallisto.^28^ The transcript-level output counts and transcripts per million (TPM) values were summed on gene level and annotated with Ensembl gene ID, gene name, and gene biotype using the R/Bioconductor packages tximport and BioMart.^29,30^

### Blood RNA sequencing data analysis

Sample processing batch effects were evaluated by principle component analysis at genome wide level (Supplementary Figure 2a) and among the intersect of the 10% genes with least variable expression in each sample processing batch (Supplementary Figure 2b). A batch effect evident in the least variant gene expression analysis was corrected using the ComBat function in the sva package in R, allocating samples with PC2 score <0 and >0 (in Supplementary Figure 2b) to separate batches^31^. PCA of the least low variance gene expression after batch correction showed no further separation of samples by processing batch (Supplementary Figure 2c). Molecular degree of perturbation (MDP) was calculated as previously described^32^. Briefly, transcripts were included if more than one sample had a TPM count above the limit of detection, and the standard deviation (SD) of TPM among uninfected controls was>0.5. The TPM values for each individual data set were then transformed to a Z score using the mean and SD for each transcript among uninfected controls used as a standard reference. The MDP of each sample/data set was then represented as the sum of all Z scores>2 divided by the total number of transcripts. Differential gene expression between data sets from individuals with co-incident infection and non-infection controls was identified using a Mann-Whitney test with false discovery rate <0.05 and absolute fold difference >1.5 (or Log_2_ 0.585). Analysis of upstream transcriptional regulation of the differentially expressed genes was performed using Ingenuity Pathway Analysis (Qiagen, Venlo, The Netherlands) and visualised as network diagram using the Force Atlas 2 algorithm in Gephi v0.9.2^33^. We depicted all statistically over-represented molecules (false discovery rate <0.05), predicted to be upstream of >2 target genes, and annotated with one of the following molecular functions: cytokine, transmembrane receptor, kinase and transcriptional regulator, representing the canonical components of molecular pathways responsible for transcriptional reprogramming in immune responses. The biological pathways represented by the upstream regulators were identified by Reactome pathway enrichment analysis using XGR^34^ as previously described^35,36^. For visualization, 20 pathway groups were identified by hierarchical clustering of Jaccard indices to quantify similarity between the gene compositions of each pathway. For each group, the pathway with the largest total number of genes was then selected to provide a representative annotation.

### Transcriptional modules

To identify co-regulated gene networks used as transcriptional modules, we calculated the average correlation coefficient for pairwise correlations of the expression levels of each group of target genes associated with predicted upstream regulators in our transcriptomic data set, and compared this to the distribution of average correlation coefficients obtained from random selection of equivalent sized groups of genes repeated 100 times. Groups of target genes with average correlation coefficients that exceeded the mean of the distribution of equivalent sized randomly selected groups by ≥2 SD (z-score ≥2) with false discovery rate <0.05 were identified as transcriptional modules representing the functional activity of the associated upstream regulator (Supplementary Figure 3). Independently derived Type 1 and Type 2 interferon inducible modules and cell-type specific transcriptional modules were described previously^35,37,38^. To derive an independent cell proliferation module, PBMC were isolated from BCG-vaccinated individuals and stimulated in vitro with 100 ng/ml purified protein derivative (PPD) for 6 days to drive proliferation of antigen specific T cells. Stimulated and unstimulated PBMC were subjected to transcriptional profiling, differential gene expression and Reactome pathway enrichment analysis as previously described^37,39^. Differentially enriched transcripts annotated to the “Cell Cycle” Reactome pathway (Supplementary table 3) were used to derive a transcriptional signature for T cell proliferation. The expression of each module was represented by the geometric the mean Log_2_ TPM value of its constituent genes.

### Data from human challenge studies

Publicly available data from previously published human viral challenge studies were downloaded from GEO (GSE73072). We calculated module scores for the STAT1 and CCND1 modules as the mean expression across all constituent genes, using log2-transformed microarray data. Only participants who developed evidence of infection following inoculation were included, as per the original study definitions^6^. The peak enrichment of STAT1 and CCND1-regulated modules for each infected individual was calculated was represented by the highest log2 TPM ratio to the mean of uninfected controls, across the time course of each data set (Supplementary Figure 6).

### Flow cytometry of peripheral blood mononuclear cells

PBMC were isolated from heparinised blood by density centrifugation using Ficoll-Hypaque Plus (GE Healthcare). PBMC were frozen in 10% DMSO (Sigma-Aldrich) in Isopropanol containers (−1°C/minute) at 5 x 10^6^ PBMC/ml in cryovials. Thawing was performed by gentle agitation at 37°C with rapid dilution in RPMI containing 10% fetal bovine serum (FBS; Sigma-Aldrich). For multiparametric flow cytometry cells were plated in 96-well round-bottomed plates (0.5-1 x 10^6^ per sample) and washed once in PBS (Phosphate buffered saline; Thermo Fisher Scientific) then stained with Blue fixable live/Dead dye (Thermo Fisher Scientific) for 20 mins at 4°C in PBS. Cells were washed again in PBS, and incubated with saturating concentrations of monoclonal antibodies (mAbs) against markers to be stained on the cell surface, diluted in 50% Brilliant violet buffer (BD biosciences) and 50% PBS for 30 min at 4°C unless stated. After surface Ab staining (Supplementary table 2) cells were resuspended in fix/perm buffer (ebiosciences, Foxp3 / Transcription Factor staining buffer kit, fix perm concentrate diluted 1:3 in fix/perm diluent) for 45-60 mins at 4°C. Cells were then washed in 1x perm buffer (10x perm buffer Foxp3 / Transcription Factor staining buffer kit diluted to 1X in ddH2O) and saturating concentrations of intranuclear targets (Ki67) were stained in 1X perm buffer for 30-45 mins 4°C. Cells were washed twice in PBS then analyzed by flow cytometry.

Samples were acquired in PBS on LSR II flow cytometer (BD biosciences) and were analyzed using FlowJo (version 10.7.1 for mac, Tree Star). Single stain controls were prepared with cells or anti-mouse IgG beads (BD biosciences). Fluorescence minus one (FMOs) were used for gating (See supplementary figure 7). For tSNE (Figure 3b) an equal number (2223 cells) of CD4 and CD8 T cells from each of the 7 control and 9 PCR+ samples were concatenated and tSNE was calculated on single cells expression values for the following markers: CD4, CD8, HLA-DR, Ki67, CD45RA, CCR7 (Iterations 1000, perplexity 30, eta 4979; KNN algorithm, Exact. Gradient algorithm, Barnes-Hut).

### T cell receptor sequencing and analysis

The α and β chains of the TCR repertoire were sequenced from all time points for which RNA was available within the first 4 weeks of the study.for all participants who were PCR+ at any time point, and for six randomly selected individuals who remained PCR- and seronegative throughout the study. The pipeline introduces unique molecular identifiers attached to individual cDNA molecules which allows correction for sequencing error PCR bias, and provides a quantitative and reproducible method of library preparation. Full details for both the experimental TCRseq library preparation and the subsequent TCR annotation (V, J and CDR3 annotation) using Decombinator V4 have been described previously^40–42^ The Decombinator software is freely available at https://github.com/innate2adaptive/Decombinator. Expanded TCRs were defined as any TCR which changed significantly between any two time points (Supplementary Figure 8). The boundaries (shown as blue dotted lines) were defined as the maximum TCR abundance which might be observed at time 2, given its abundance at time 1, assuming Poisson distribution of counts with p < 0.0001, to give a false discovery rate of <1 in 1000. TCR abundances are normalised for total number of TCRs sequenced in each sample, and expressed as counts/million. MAIT TCRs were defined as any TCR alpha containing TRAV1-2 paired with TRAJ12, TRA20 or TRAJ33. iNKT TCRs were defined as TCRs containing TRAV24 paired with TRAJ18. The VDJdb database (https://vdjdb.cdr3.net/about) was searched for any TCR annotated for CMV, EBV or SARS-Cov-2. TCRs annotated for multiple antigens were excluded. This set of antigen-associated TCRs were then compared to our set of expanded TCRs defined as described above.

### Data sharing statement

Applications for access to the individual participant de-identified data (including data dictionaries) and samples can be made to the access committee via an online application *https://covid-consortium.com/application-for-samples/*. Each application will be reviewed, with decisions to approve or reject an application for access made on the basis of (i) accordance with participant consent and alignment to the study objectives (ii) evidence for the capability of the applicant to undertake the specified research and (iii) availability of the requested samples. The use of all samples and data will be limited to the approved application for access and stipulated in the material and data transfer agreements between participating sites and investigators requesting access.

### Data and materials availability

RNAseq data, TCR sequencing data and associated essential metadata will be made publicly available at time of peer-reviewed publication.

## Supporting information

Consortium investigator list

## Data Availability

Open access to RNAseq and TCRseq data and associated essential metadata will be made available after peer-reviewed publication. Applications for access to the individual participant de-identified data (including data dictionaries) and samples can be made to the access committee via an online application https://covid-consortium.com/application-for-samples/. Each application will be reviewed, with decisions to approve or reject an application for access made on the basis of (i) accordance with participant consent and alignment to the study objectives (ii) evidence for the capability of the applicant to undertake the specified research and (iii) availability of the requested samples. The use of all samples and data will be limited to the approved application for access and stipulated in the material and data transfer agreements between participating sites and investigators requesting access.

## Acknowledgements

Funding for COVIDsortium was donated by individuals, charitable Trusts, and corporations including Goldman Sachs, Citadel and Citadel Securities, The Guy Foundation, GW Pharmaceuticals, Kusuma Trust, and Jagclif Charitable Trust, and enabled by Barts Charity with support from UCLH Charity. RKG is funded by National Institute for Health Research (DRF-2018-11-ST2-004). JCM, CM, and TAT are directly and indirectly supported by the University College London Hospitals (UCLH) and Barts NIHR Biomedical Research Centres, and through the British Heart Foundation (BHF) Accelerator Award (AA/18/6/34223). TAT is funded by a BHF Intermediate Research Fellowship (FS/19/35/34374). MN is supported by the Wellcome Trust (207511/Z/17/Z) and by NIHR Biomedical Research Funding to UCL and UCLH. RJB/DMA are supported by UKRI/MRC Newton (MR/S019553/1, MR/R02622X/1, MR/V036939/1MR/S019553/1, and MR/R02622X/1), NIHR Imperial Biomedical Research Centre (BRC):ITMAT, Cystic Fibrosis Trust SRC, and Horizon 2020 Marie Curie Actions. MKM is supported by the UKRI/NIHR UK-CIC grant, a Wellcome Trust Investigator Award (214191/Z/18/Z), and a CRUK Immunology grant (26603) AM is supported by Rosetrees Trust, The John Black Charitable Foundation, and Medical College of St Bartholomew’s Hospital Trust.

