## Supplementary Tables & Figs for "Non-severe SARS-CoV-2 infection is characterised by very early T cell proliferation independent of type 1 interferon responses and distinct from other acute respiratory viruses"

***Supplementary table 1. Baseline characteristics of the study cohort.***

| Characteristic | Overall, N = 96 | Cases, N = 41[1] | Controls, N = 55[1] |
|---|---|---|---|
| Age | 36 (27, 47) | 36 (28, 46) | 36 (26, 50) |
| Sex | | | |
| Female | 69 (72%) | 28 (68%) | 41 (75%) |
| Male | 27 (28%) | 13 (32%) | 14 (25%) |
| Ethnicity | | | |
| White | 66 (69%) | 25 (62%) | 41 (75%) |
| Black | 6 (6·3%) | 5 (12%) | 1 (1·8%) |
| Asian | 18 (19%) | 9 (22%) | 9 (16%) |
| Other | 5 (5·3%) | 1 (2·5%) | 4 (7·3%) |
| Unknown | 1 | 1 | 0 |
| Samples | | | |
| 1 | 67 (70%) | 12 (29%) | 55 (100%) |
| 2 | 7 (7·3%) | 7 (17%) | 0 (0%) |
| 3 | 6 (6·2%) | 6 (15%) | 0 (0%) |
| 4 | 10 (10%) | 10 (24%) | 0 (0%) |
| 5 | 6 (6·2%) | 6 (15%) | 0 (0%) |
| Case-defining symptoms | 36 (38%) | 31 (76%) | 5 (9·1%) |

[1]Statistics presented: median (IQR); n (%)

***Supplementary table 2. Flow cytometry antibody reagents***

| Target | Fluorochrome | Clone | Manufacturer | Catalogue no |
|---|---|---|---|---|
| TCR Va7.2 | BV785 | 3C10 | Biolegend | 351722 |
| CCR7 | FITC | 150503 | BD | 561271 |
| CD45RA | BV711 | HI100 | BD | 563733 |
| CD4 | BUV395 | SK3 | BD | 563550 |
| CD56 | Pe-Dazzle594 | QA17A16 | Biolegend | 392410 |
| HLA-DR | V500 | G46-6 | BD | 561224 |
| CD3 | BUV805 | UCHT1 | BD | 612895 |
| CD161 | Pe | 191B8 | Miltenyi | 130-092-677 |
| CD8a | AlexaFluor700 | RPA-T8 | Biolegend | 301028 |
| Ki67 | Pe-Cy7 | 20Raj1 | ThermoFisher | 25-5699-42 |
| OX40 | AlexFluor647 | Ber-ACT35 | Biolegend | 350018 |
| PD-1 | BB700 | EH12.1 | BD | 566460 |
| CD69 | BV605 | FN50 | Biolegend | 310938 |
| CD71 | APC-Cy7 | CY1G4 | Biolegend | 334110 |
| 4-1BB | BV421 | 4B4-1 | Biolegend | 309820 |

***Supplementary Table 3. Genes comprising the T cell proliferation module derived from BCG stimulated PBMC.***

| Gene symbol | Ensembl gene ID |
|---|---|
| AURKA | ENSG00000087586 |
| BUB1 | ENSG00000169679 |
| BUB1B | ENSG00000156970 |
| CCNA2 | ENSG00000145386 |
| CCNB1 | ENSG00000134057 |
| CCNB2 | ENSG00000157456 |
| CDCA5 | ENSG00000146670 |
| CDCA8 | ENSG00000134690 |
| CDK1 | ENSG00000170312 |
| CDK6 | ENSG00000105810 |
| CDKN2C | ENSG00000123080 |
| CDT1 | ENSG00000167513 |
| CENPM | ENSG00000100162 |
| CENPN | ENSG00000166451 |
| GINS2 | ENSG00000131153 |
| H2AC14 | ENSG00000276368 |
| H2AFX | ENSG00000188486 |
| H3C10 | ENSG00000278828 |
| H3C12 | ENSG00000197153 |
| H3C15 | ENSG00000203852 |
| H3C4 | ENSG00000197409 |
| HIST2H3D | ENSG00000183598 |
| KIF2C | ENSG00000142945 |
| LMNB1 | ENSG00000113368 |
| MLF1IP | ENSG00000151725 |
| NDC80 | ENSG00000080986 |
| PCNA | ENSG00000132646 |
| PTTG1 | ENSG00000164611 |
| SMC4 | ENSG00000113810 |

### Supplementary Figures

***Supplementary Figure 1. Study consort diagram***

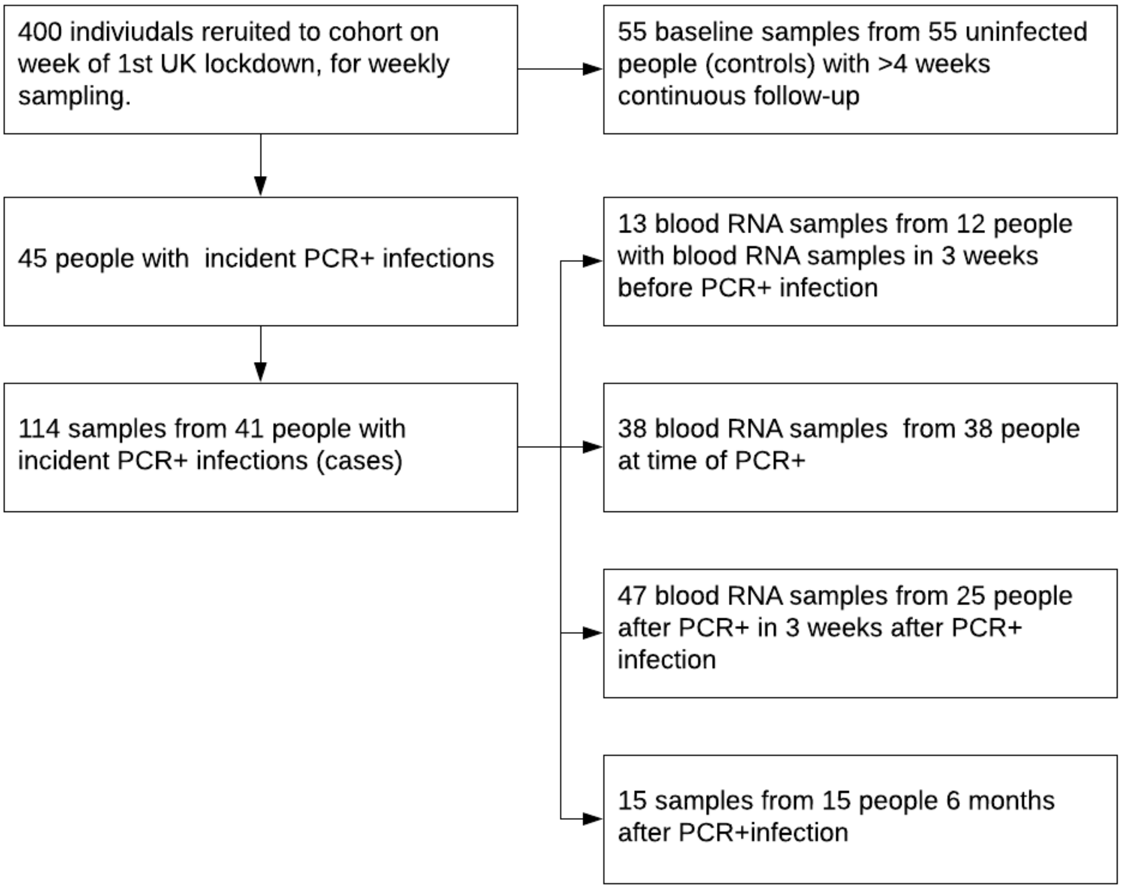

**(A)** Consort diagram of nested case-control study. **(B)** Molecular degree of perturbation of blood transcriptomes from participants with co-incident PCR+ve SARS-CoV-2 infection with and without contemporary case defining symptoms compared to non-infected controls.

***Supplementary Figure 2. Principal component analysis of blood transcriptional profiles stratified by sample processing batch.***

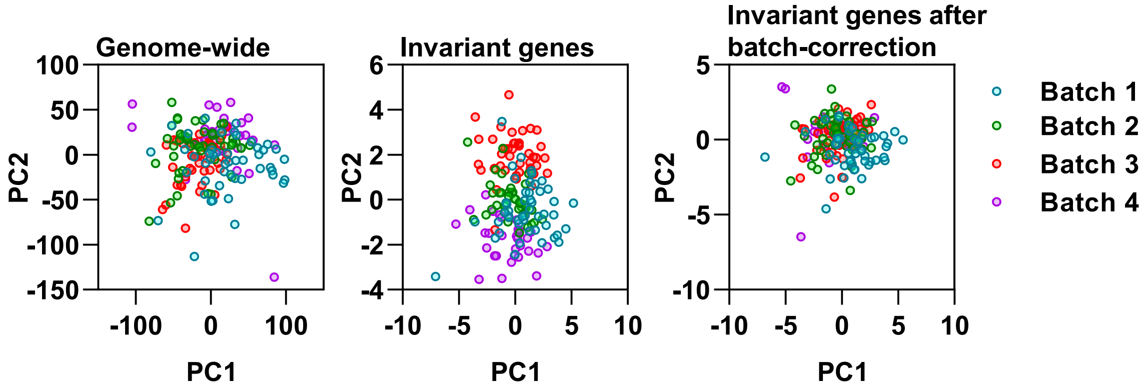

***Supplementary Figure 3. Blood transcriptomic perturbation in symptomatic and asymptomatic SARS-CoV-2 infection.***

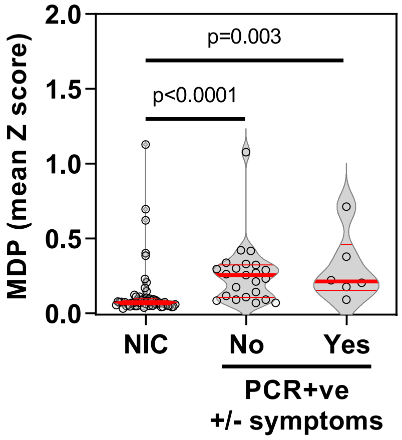

Molecular degree of perturbation (MDP) in blood transcriptomes for each individual expressed as the mean of genome-wide standard deviations (Z scores) >2 from the mean of non-infection controls (NIC), among samples from non-infection controls and samples from individuals with co-incident PCR+ve infection with (Yes) or without (No) case-defining symptoms. Individual data points shown with violin plots depicting median, IQR and frequency distributions. P values derived from Mann-Whitney tests.

***Supplementary Figure 4. Identification of co-regulated modules in blood transcriptomic data***

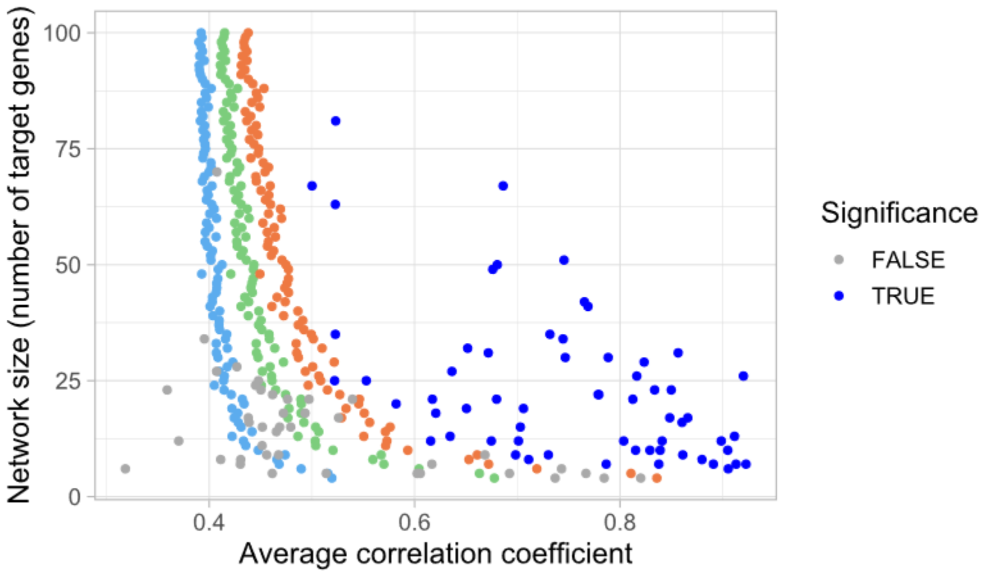

Z-scores derived from the distribution of average correlation coefficients obtained from 100 iterations of randomly selecting groups of genes from blood transcriptomic data are shown in light blue (z-score=1), green (z-score=2) and orange (z-score=3). Average correlation coefficients of molecular networks associated with upstream regulators of differentially expressed genes between individuals with incident SARS-CoV-2 infection and noninfection controls are shown in dark blue (z-score ≥2, false discovery rate ≤0.05) and grey (z-score ≤2 +/- FDR ≥0.05) compared to equivalent sized random gene networks.

***Supplementary Figure 5. Bioinformatic analysis of modules derived from upstream regulator analysis of differentially expressed genes in blood transcriptome associated with SARS-CoV-2 infection.***

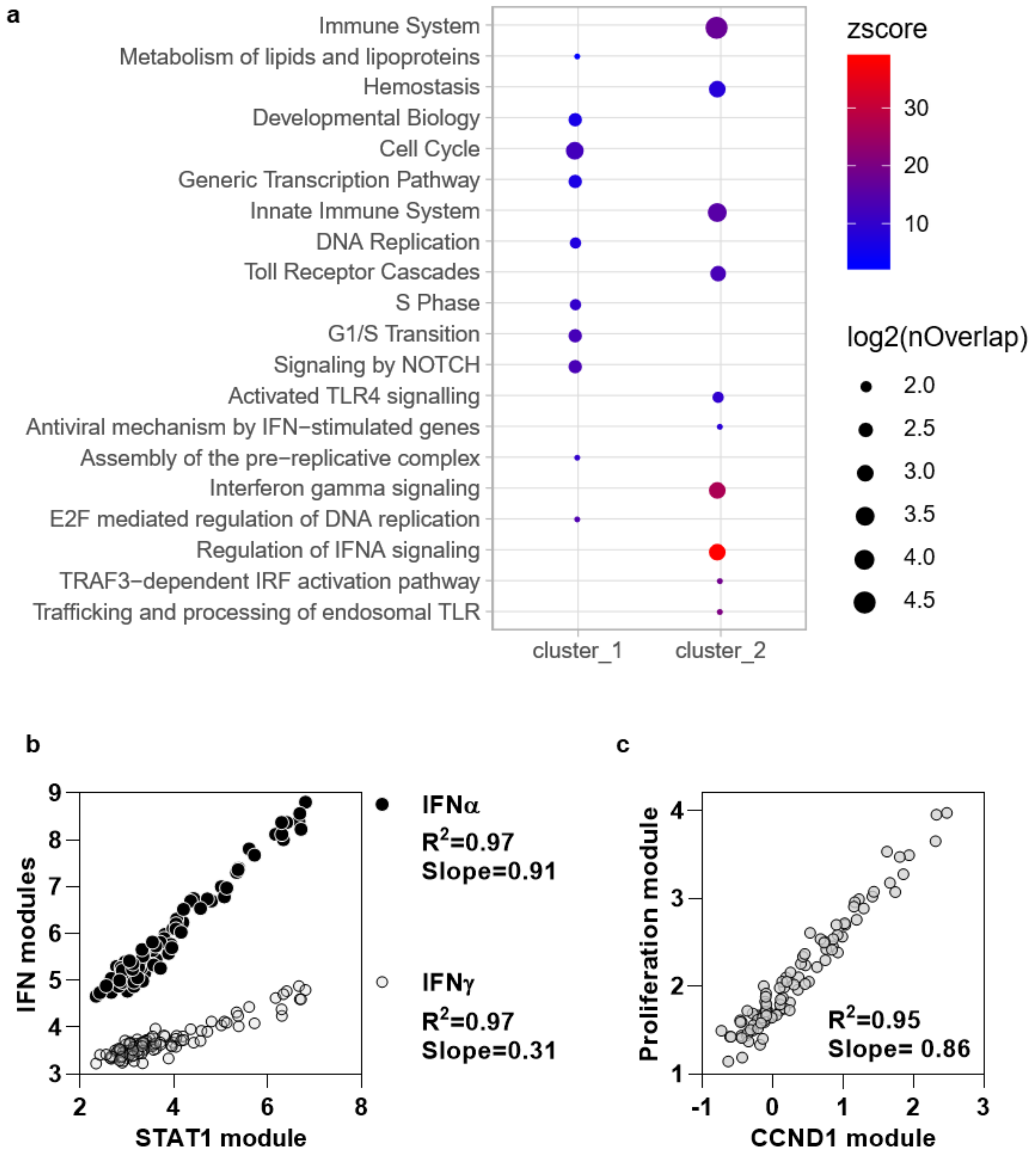

**(A)** Enrichment of Reactome pathways among the two clusters of predicted upstream regulators associated with co-incident infection (depicted main manuscript Figure 1 and listed Supplementary Data File 1). Node size represents number of upstream regulators associated with each pathway and statistical enrichment represented by Z score. **(B)** Correlation of STAT1-regulated transcriptional module with interferon (IFN)α and IFNγ modules, and **(C)** correlation of CCND1-regulated transcriptional module with independently derived proliferation module in all time points (-3 to +3 weeks) from individuals with SARS-CoV-2 infection.

***Supplementary Figure 6. STAT1 and CCND1 regulated module expression over time in blood transcriptomic data from individuals experimentally infected with acute respiratory viruses.***

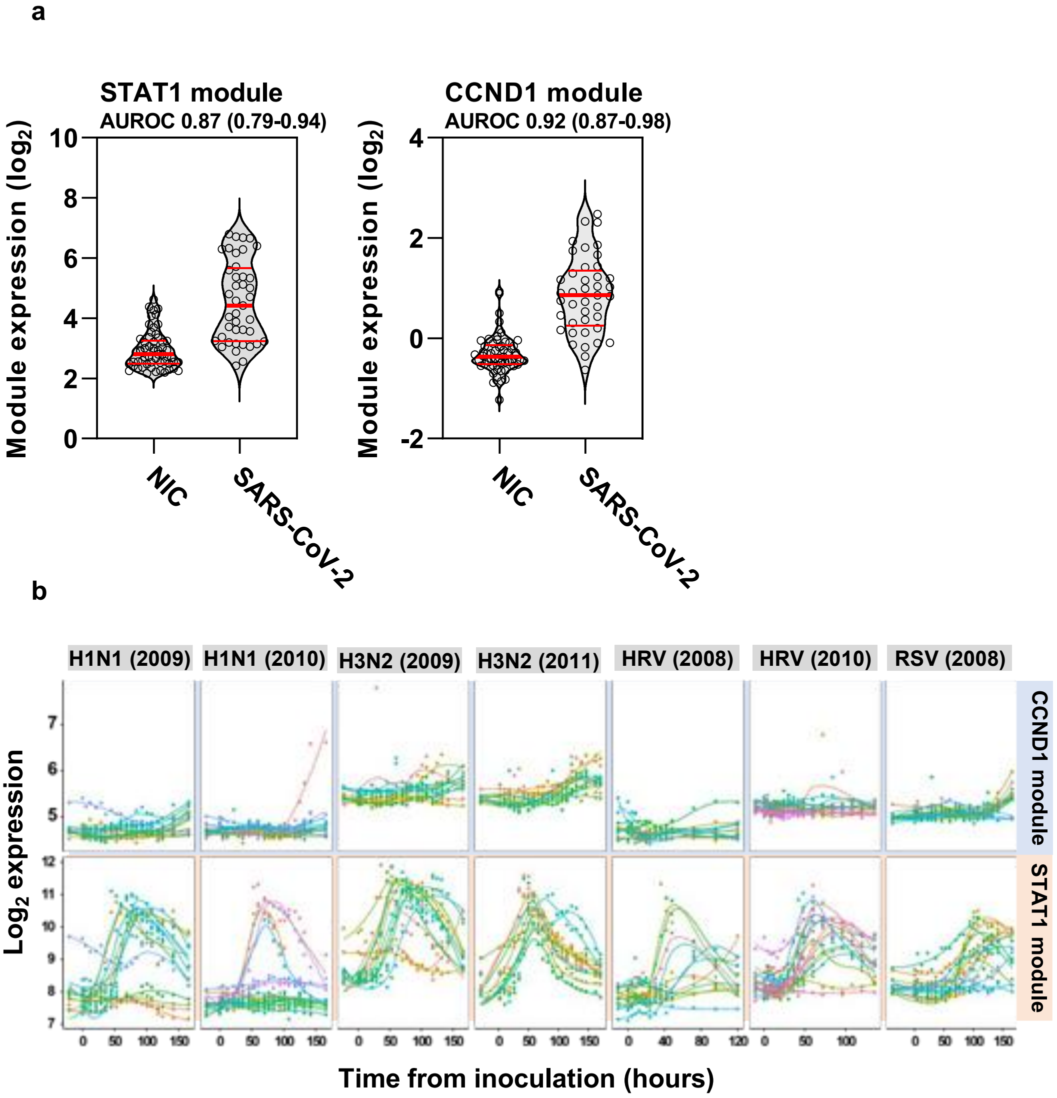

**(A)** Comparison of peak STAT-1 and CCND1-regulated module expression in samples from individuals with SARS-CoV-2 infection and non-infection controls showing area under the receiver operating curve ±95% confidence intervals for these modules to discriminate between the two groups. **(B)** Expression scores for STAT1 and CCND1 modules in publicly available data from seven human viral challenge studies (GSE73072). Analysis restricted to participants with evidence of infection following inoculation, as per original study definition (total n=92 participants). Colours represent individual participants with longitudinal sampling. Each column represents a different challenge study, with the virus and study year shown in the column headers. HRV = human rhinovirus; RSV = respiratory syncytial virus.

***Supplementary Figure 7. Multiparametric flow cytometry of PBMC samples from individuals with co-incident SARS-CoV-2 infection and uninfected controls.***

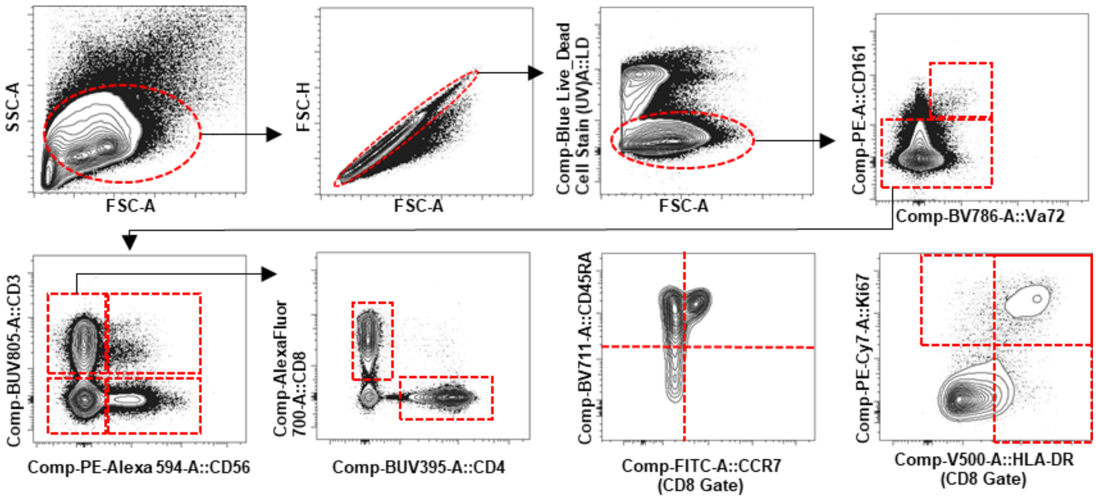

**(A)** Example gating strategy for flow cytometric data showing gating for; lymphocytes, singlets, live cells (fixable live dead-), MAIT cells (CD161++ TCR Va7.2+) and non-MAIT cells, CD3+CD56- T cells, CD3+CD56+ NKT cells, CD3-CD56- lymphocytes, CD3-CD56+ NK cells, CD4+ and CD8+ T cells, CD45RA vs. CCR7 co-staining, and HLA-DR vs. Ki67 co-staining.

***Supplementary Figure 8. Frequency of selected lymphocytes and their activation/proliferative state.***

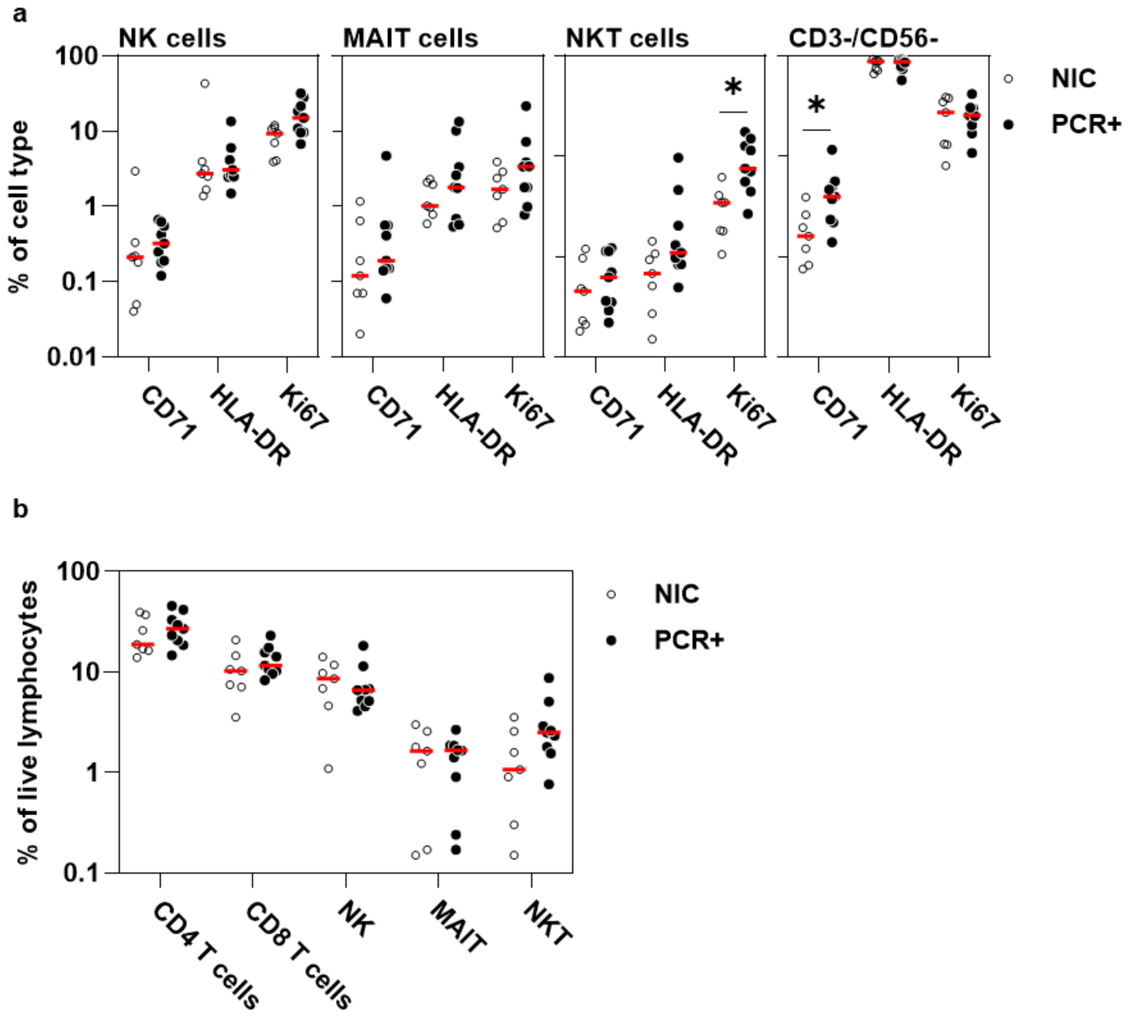

**(A)** Frequency of non-T cell lymphocyte subsets expressing selected activation markers (CD71 and HLA-DR) and the cell proliferation marker (Ki67), and **(B)** frequency of selected lymphocyte subsets among participants with co-incident infection (PCR+) and non-infection controls (NIC). Data points with median values (red). (*p<0.05 by Mann-Whitney Test for each group compared to NIC).

***Supplementary Figure 9. TCR sequence analysis***

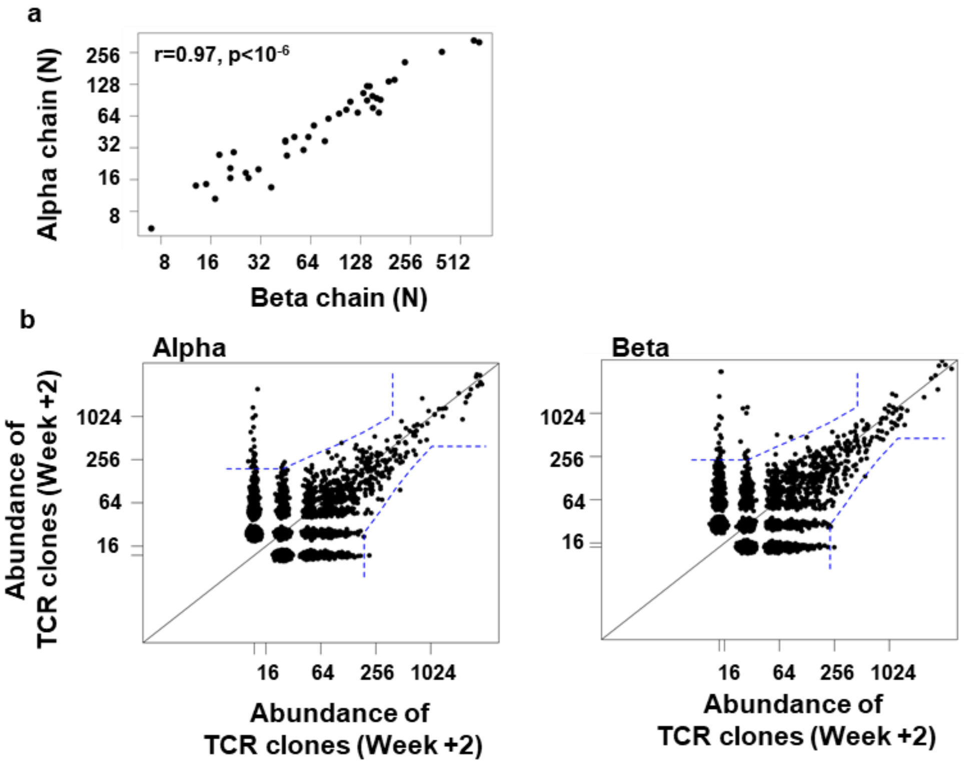

**(A)** Correlation of numbers of alpha and beta chain TCR sequences per sample. **(B)** Representative examples of identification of statistically expanded alpha, and beta chain TCR sequences by comparison of abundance of individual TCR sequences between two time points (blue dashed boundaries represent the thresholds for abundance of TCR sequences with false discovery rate of <1 in 1000).

***Supplementary Figure 10. Expansion of TCR beta chain sequences and abundance of MAIT/iNKT cell sequences in SARS-CoV-2 infection***

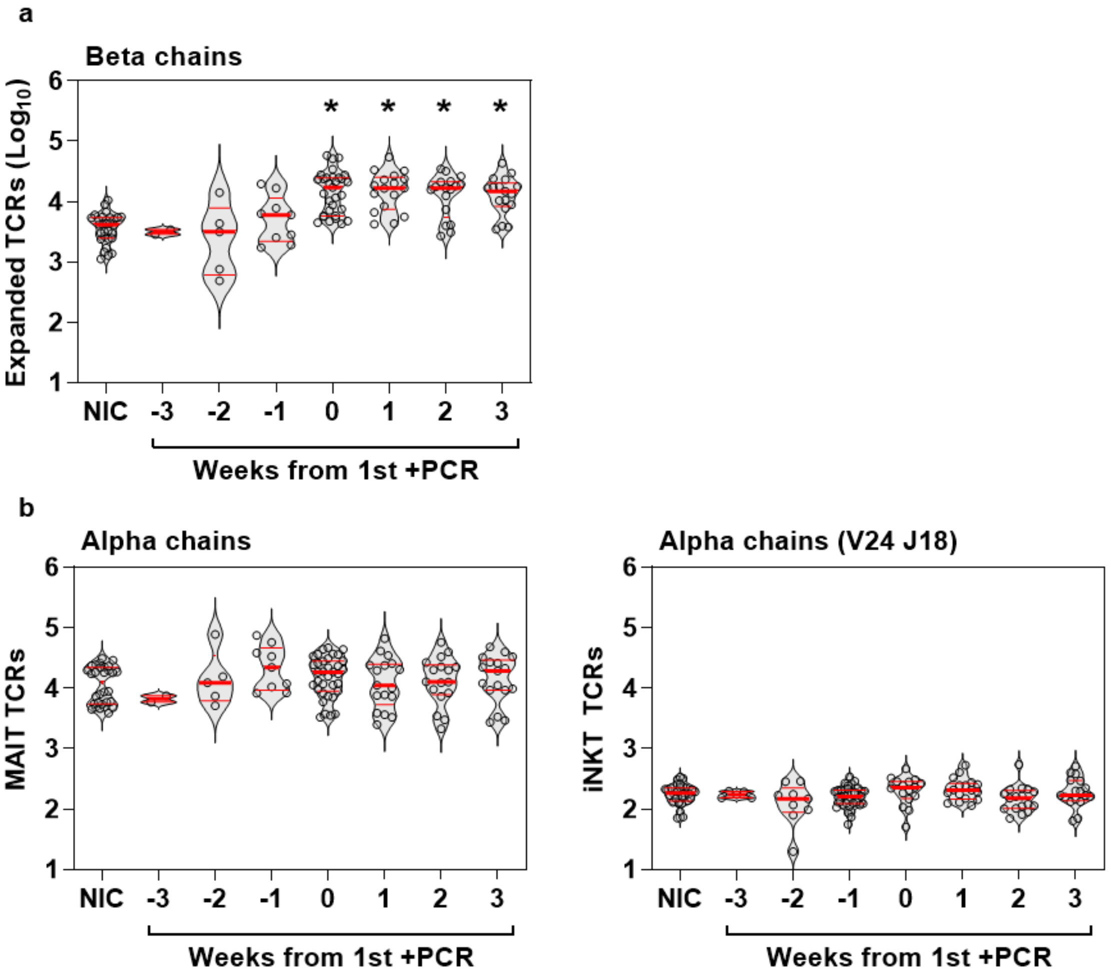

**(A)** Enumeration of expanded TCR alpha chain sequences (per million total sequences) in non-infection controls and samples from infected individuals stratified by time from first positive PCR. **(B)** Abundance of MAIT and iNKT cell associated alpha chain sequence in non-infection controls and samples from infected individuals stratified by time from first positive PCR. Individual data points shown with violin plots depicting median, IQR and frequency distributions. (*FDR<0.05 by Kruskal-Wallis Test for each group compared to NIC).
