## Supplementary material for "Non-severe SARS-CoV-2 infection is characterised by very early T cell proliferation independent of type 1 interferon responses and distinct from other acute respiratory viruses": Consortium investigator list

**COVIDsortium investigators (alphabetical order)**

| |
|---|
| Hakam Abbass |
| Aderonke Abiodun |
| Mashael Alfarih |
| Zoe Alldis |
| Daniel M Altmann |
| Mervyn Andiapen |
| Jessica Artico |
| Joao Augusto |
| Georgina L Baca |
| Anish Bhuva |
| Alex Boulter |
| Ruth Bowles |
| Rosemary J Boyton |
| Olivia Bracken |
| Timothy Brooks |
| Natalie Bullock |
| Gabriella Captur |
| Benny Chain |
| Nicola Champion |
| Carmen Chan |
| Jorge Couto de Sousa |
| Xose Couto-Parada |
| Marie-Teresa Cutino-Moguel |
| Rhodri H Davies |
| Keenan Dieobi-Anene |
| Karen Feehan |
| Malcolm Finlay |
| Marianna Fontana |
| Nasim Forooghi |
| Joseph M Gibbons |
| Derek Gilroy |
| Peter Griffiths |
| Rishi K Gupta |
| Matt Hamblin |
| Lauren M Hickling |
| Aroon D Hingorani |
| Lee Howes |
| Ivie Itua |
| Victor Jardim |
| Melanie Jensen |
| Meleri Jones |
| George Joy |
| Vikas Kapil |
| Jonathan Lambourne |
| WY Jason Lee |

| |
|---|
| Mala K Maini |
| Vineela Mandadapu |
| Charlotte Manisty |
| Aine McKnight |
| Katia Menacho Medina |
| Celina Mfuko |
| Oliver Mitchelmore |
| James C Moon |
| Mahdad Noursadeghi |
| Ben O’Brien |
| Ben Ollivere |
| Corinna Pade |
| Susana Palma |
| Kush Patel |
| Ruth Parker |
| Brian Piniera |
| Alicja Rapala |
| Amy Richards |
| Mathew Robathan |
| Genine Sambile |
| Amanda Semper |
| Andreas Seraphim |
| Angelique Smit |
| Michelle Sugimoto |
| George D Thornton |
| Thomas A. Treibel |
| Arthur Tucker |
| Ana Valdes |
| Jessry Veerapen |
| Mohit Vijayakumar |
| Timothy Warner |
| Sophie Welch |
| Dylan Williams |
| Theresa Wodehouse |
| Lucinda Wynne |
| Dan Zahedi |
